# Are health indicators able to describe the ability to cope of Health Systems with COVID-19 epidemic?

**DOI:** 10.1101/2020.06.13.20130146

**Authors:** Silvana Castaldi, Ester Luconi, Bruno Alessandro Rivieccio, Patrizia Boracchi, Giuseppe Marano, Elena Pariani, Luisa Romanò, Francesco Auxilia, Federica Nicolussi, Alessandra Micheletti, Giancarlo Manzi, Silvia Salini, Massimo Galli, Elia Biganzoli

**Affiliations:** Department of Biomedical Sciences for Health University of Milan, Italy; Fondazione IRCCS Ca’ Granda Ospedale Maggiore di Milano, Italy; Department of Clinical Sciences and Community Health & DSRC, University of Milan, Italy; Niguarda Hospital – AREU – Regional Emergency Service Agency – Milano, Italy; Department of Economics, Management and Quantitative Methods & DSRC, University of Milan, Italy; Department of Environmental Science and Policy, University of Milan, Italy; Department of Biomedical and Clinical Sciences “L. Sacco”, University of Milan, Italy

**Keywords:** COVID-19, epidemic, health indicators, health systems, Italy

## Abstract

The Coronavirus Disease 19 epidemic is an infectious disease which was declared as a pandemic and hit all the Countries, all over the world, from the beginning of the year 2020.

Despite the emergency vigilance plans, in all the Countries, Health Systems experienced a different ratio of lethality, admissions to intensive care units and managing quarantine of positive patients.

The aim of this study is to investigate if some health indicators might have been useful to understand the capacity of Italian National Health Service to manage the COVID 19 epidemic.

We will compare data in two different Italian regions in the Northern part of Italy (Lombardy and Veneto) with the national data to understand if different health strategies might be significant to explain different patterns of COVID 19 epidemic in Italy.

The two regions have two different health policies to face CoViD-2019 epidemic.

To face epidemic like this one the answer should be outside hospitals but this means to have general practitioners well-trained and enough healthcare personnel working outside hospitals.

## Introduction

Planning health services (HSs) is complex. It requires a rigorous, evidence-based approach to improving high quality services to meet the future health needs of the population (1).

This goal should be achieved through the provision of efficient and effective HS, considering available resources and balancing hospital and local services (2).

Health planning should be future oriented and able to support organizations, to be better prepared to address emerging health threats and, for this reason, it needs to develop emergency plans to assure operational readiness to respond to emergencies (2).

Public health is constantly threatened by a wide range of hazards. Despite measures to prevent them, emergencies of varying types, scales and consequences still occur and must find HS ready to manage them.

The Coronavirus Disease 19 (COVID 19) epidemic is an infectious disease which was declared as a pandemic and hit all the Countries, all over the world, from the beginning of the year 2020 (3).

Despite the emergency vigilance plans, in all the Countries, HSs experienced a different ratio of lethality, admissions to intensive care units (ICUs), and managing quarantine of positive patients (3,4).

We might evaluate these differences as a proxy of distinct management decisions at both national and local level, because health data related to COVID-19 infection might change with its different healthcare management strategies, for example:

- Observed lethality ratio might change in relation of the number of the swabs performed (only with the application of the swabs we can be sure about the positive cases and the correct diagnosis).
- Some health authorities decided treating COVID-19 positive patients at home and some others in hospital. We should indeed consider that to treat COVID-19 positive patients in hospital might expose healthcare workers to the infection, moreover quarantine or home isolation of a-/pauci-symptomatic patients could leave hospital beds free for more severe cases (5).

The aim of this study is to investigate if the health indicators took into consideration might be useful to understand the capacity of Italian HS to manage the COVID 19 epidemic.

We will compare data in two different Italian regions in the Northern part of Italy (Lombardy and Veneto) with the national data to understand if different HS strategies might be significant to explain different patterns of COVID 19 epidemic in Italy.

The considered indicators are not good to plan preventive measures, but they are quite good measures to estimate the burden of the disease in this pandemic situation.

## Materials and Methods

Data about the spread of the coronavirus in Italy were provided by “Dipartimento della Protezione Civile – Presidenza del Consiglio dei Ministri” (http://www.protezionecivile.gov.it/it); in particular the following information were update daily at 18:30 (after the Head of Department press conference): national situation, regional situation and provinces situation.

We obtained the data from the site of GitHub in which there is a repository of the data of “Dipartimento della Protezione Civile” (https://github.com/pcm-dpc/COVID-19).

For this work we took into consideration national data from 24/02/2020 to 12/04/2020 (so 49 days in total) and in particular those variables:

- deaths
- symptomatic hospitalized patients
- patients in ICUs
- currently infected patients
- total cases (currently positive patients, healed patients and deaths)
- patients in quarantine
- discharged or healed patients

For a particular day those variables are presented as a cumulate of the values of previous days. We tried to model the dynamic evolution change of those indicators during the considered period:

- The ratio between discharged or healed patients and total cases
- Case fatality ratio (“observed lethality”)
- The ratio between currently infected patients and total cases
- The ratio between ICU patients and currently infected patients
- The ratio between quarantined patients and currently infected patients
- The ratio between symptomatic hospitalized patients and currently infected patients

For each day, data were available in form of aggregated daily counts and only the numerator and denominator to calculate the endpoints of interest for each analysis were used.

A logistic regression model was used where the response variable was the proportion of subjects with the endpoint above reported and the independent variable was the time starting from 24/02/2020 (beginning of the spread of the coronavirus) and the day of 12/04/2020.

In the model, the denominator used to calculate the outcome of interest was added to the model as weight; for example, for the ratio between discharged/healed patients and total cases, the denominator was the number of total cases.

The shape of the time trend was modelled including restricted cubic spline functions.

Our aim was simply to obtain a smoothed shape of the trends, so we avoid to define knots positions.

Knots were placed according to a standard procedure, suggested by Harrell (6); in particular given the linearity constraint, the first knot is placed at 0.025 quantile and the last at 0.975 quantile of the time distribution. In fact, restricted cubic splines, depend on the number of knots but are robust to the exact knots position.

Separate models were performed for Italy, Lombardy and Veneto for each indicator.

In order to choose the final model representing the smoothed shape of the trends the following procedure was applied:

1. Models with spline from 3 to 10 knots were performed, because we decided to have a maximum of one knot every five days.
2. For each model AIC (Akaike Information Criterion) was reported and the estimated trend of the indicator over time was graphically examined in relationship to the observed data.
3. Among all the models that one with the lower value of AIC was considered.

The following splines were chosen for each index:

- The ratio between discharged or healed patients and total cases: for Italy, Lombardy and Veneto we chose a spline with 10 knots.
- Case fatality ratio (“observed lethality”): for Italy we chose a spline with 8 knots, for Lombardy we chose a spline with 10 knots, while for Veneto we chose a spline with 5 knots.
- The ratio between currently infected patients and total cases: for Italy, Lombardy and Veneto we chose a spline with 10 knots.
- The ratio between ICUs patients and currently infected patients: for Italy we chose a spline with 10 knots, for Lombardy we chose a spline with 7 knots and for Veneto we chose a spline with 9 knots.
- The ratio between quarantined patients and currently infected patients: for Italy and for Lombardy we chose a spline with 10 knots, while for Veneto we chose a spline with 9 knots.
- The ratio between symptomatic hospitalized patients and currently infected patients: for Italy and Lombardy we chose a spline with 10 knots, while for Veneto we chose a spline with 8 knots.

As the estimated regression coefficients for the cubic splines terms are not directly interpretable, results are reported only by the graphical estimated trend.

In some smoothed estimates, the goodness of fit for the first days (in general for the first 10 days) seems to be not satisfactory, but this is because of the low number of events in that period.

## Results

Fig. 1 shows the ratio between discharged or healed patients and total cases.

**Fig. 1.**
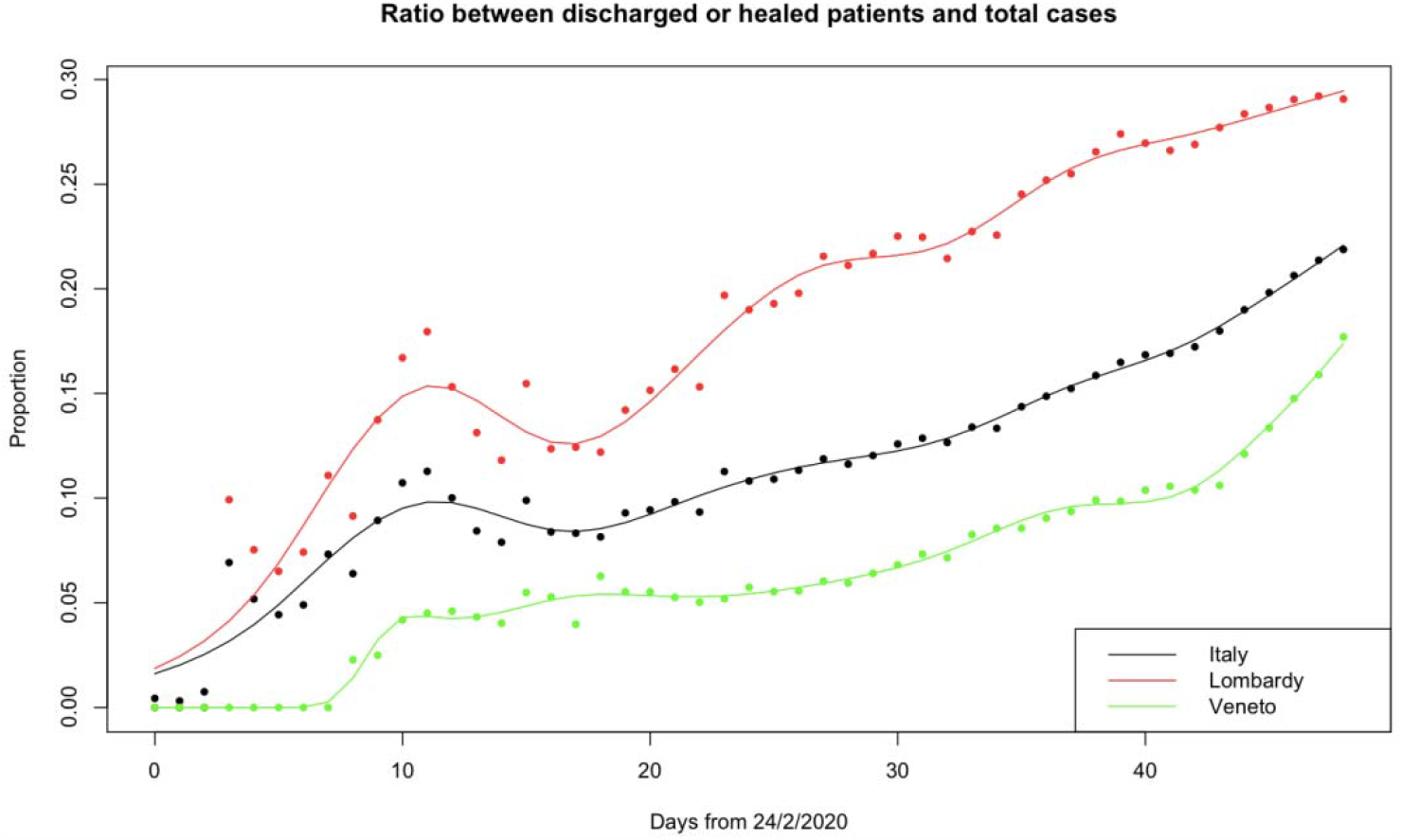
Ratio between discharged or healed patients and total cases.

The trend of the Lombardy is over the trend of Italy and Veneto, but all the three trends show an increase during the time of our observation as it is expected in an epidemic.

In Veneto the trend is underlying that of Lombardy, because of the different strategies adopted by the regions: in Veneto we had less hospitalization (and so less discharged patients) because hospitalization was decided only for patients with critical condition; this strategy probably had a positive impact for the healed patients.

Fig. 2 shows the case fatality ratio (“observed lethality”)

**Fig. 2.**
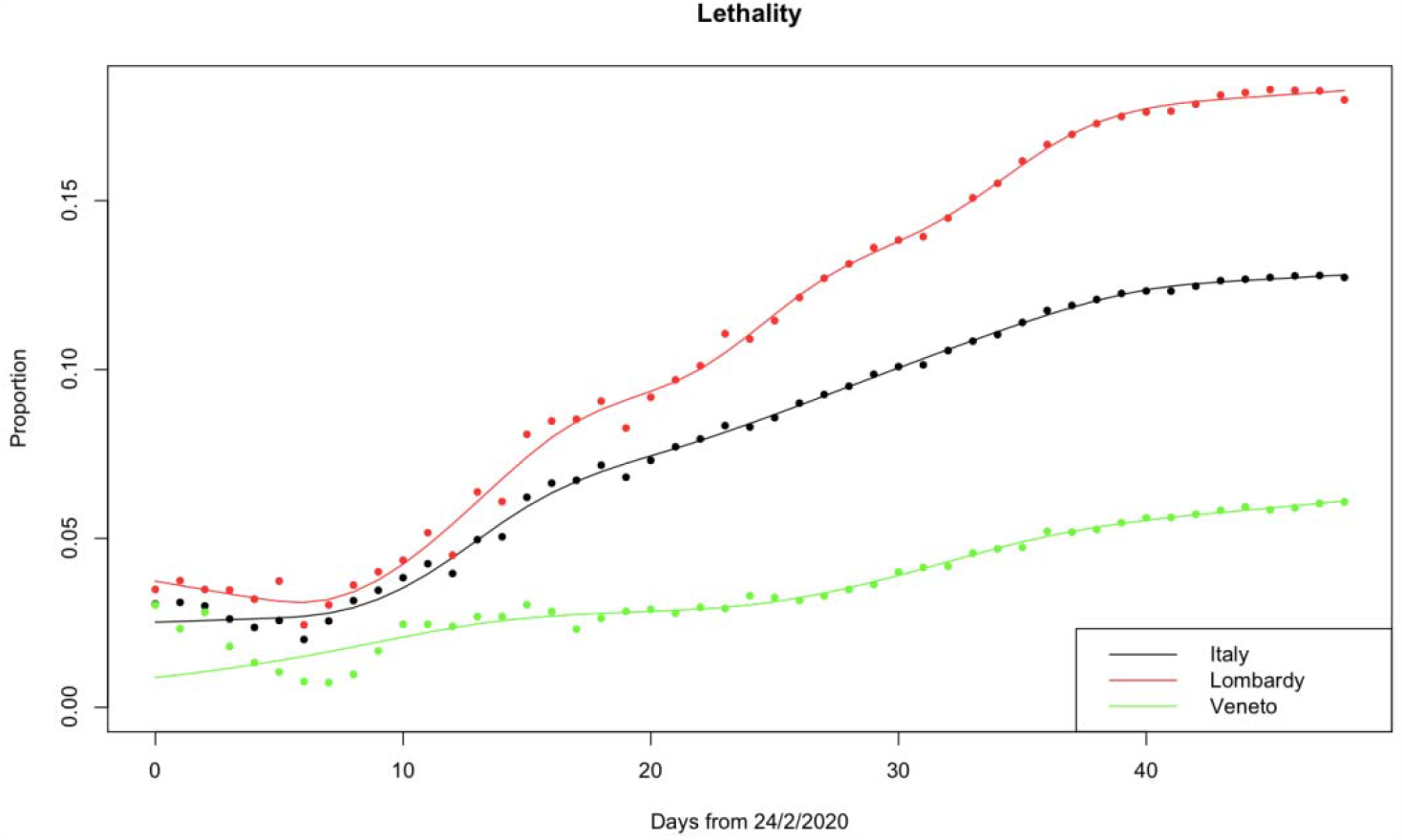
Case fatality ratio (“observed lethality”)

The lethality was very high in Lombardy. This may be explained by the fact that epidemic started in this region for the intensive commercial traffic with China.

The beginning of the epidemic was misunderstood probably because it happened during the influenza season.

There are also other explanations for this:

- With a very high number of admissions in hospital it is possible that all the deaths occurred in hospital were considered caused by COVID 19 which it might have been only a comorbidity.
- The high admission rate might be a measure of the inadequate answer for the cure of the disease outside hospitals.
- The high lethality rate in Lombardy might be explained by the high number of deaths in old people guested in residential homes.
- It might be a bias the low lethality rate in Veneto only because people died at home with no swab performed and no identification of the positive condition for COVID 19.

We may distinguish between case fatality ratio (“observed lethality”) vs. infected fatality ratio (“true lethality”).

One can argue that in Lombardy lethality “appears” so high because of the underestimation of the number of real total cases, due to the low number of swabs performed and the consequent underestimation of the denominator. It must be considered that also the numerator of the fraction is certainly underestimated, since many people died at home (or even in hospitals, at least at the beginning of epidemic) without a certain diagnosis.

The red areas located in Bergamo and Brescia, which are in Lombardy, might have contributed to this high number of deaths.

The rapid increase of the number of deaths put the Government to decide for some restriction measures which became the national lockdown on the 9^th^ of March (7).

Fig. 3 shows the ratio between currently infected patients and total cases

**Fig. 3.**
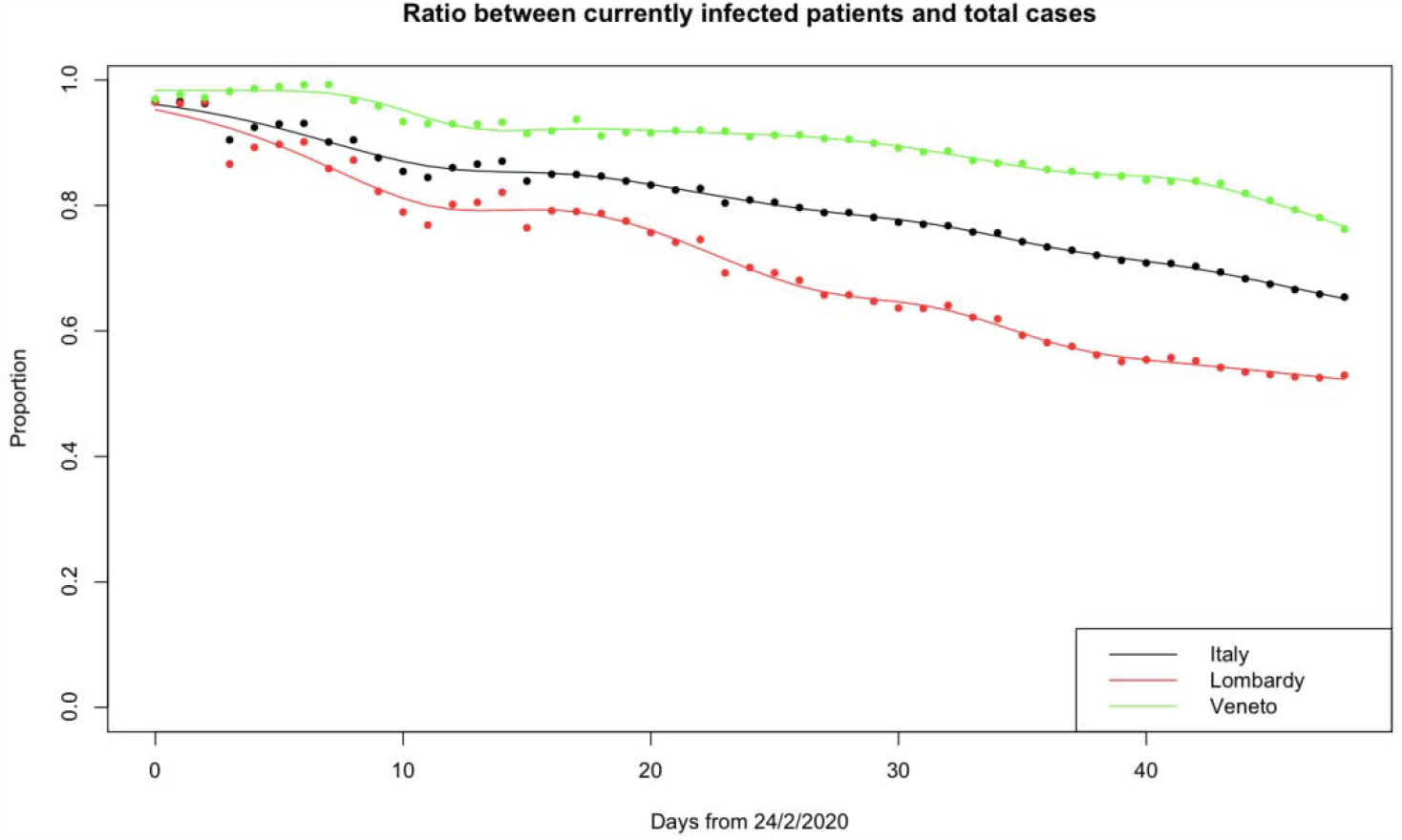
Ratio between currently infected patients and total cases

The trends, as we expected, reflect the performance of an epidemic: the number of positive falls because patients recover and thank to the lockdown measure. The numbers of Veneto are higher only because it performed much more swabs.

Fig. 4 shows the ratio between ICUs patients and currently infected patients

**Fig. 4.**
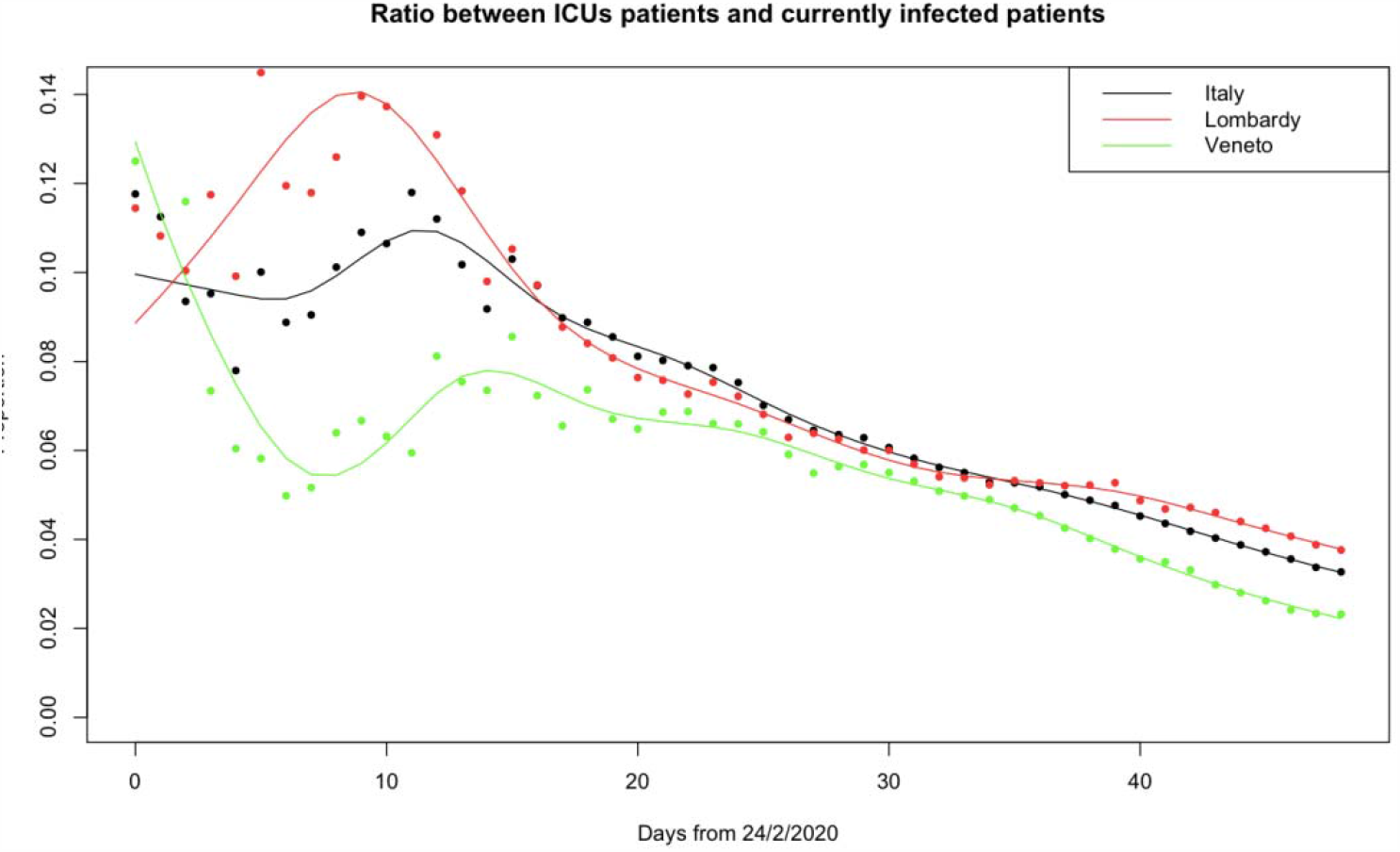
Ratio between ICUs patients and currently infected patients

The intensive use of hospital beds is evident in the figure for Lombardy while in Veneto the use of ICUs is postponed of some days and devoted only to the more severe patients. Lombardy faced the highest number of cases at the beginning of the epidemic when the way to cure these patients was not assessed and the intensive care would have been the best therapeutic option.

Fig. 5 shows the ratio between quarantined patients and currently infected patients

**Fig. 5.**
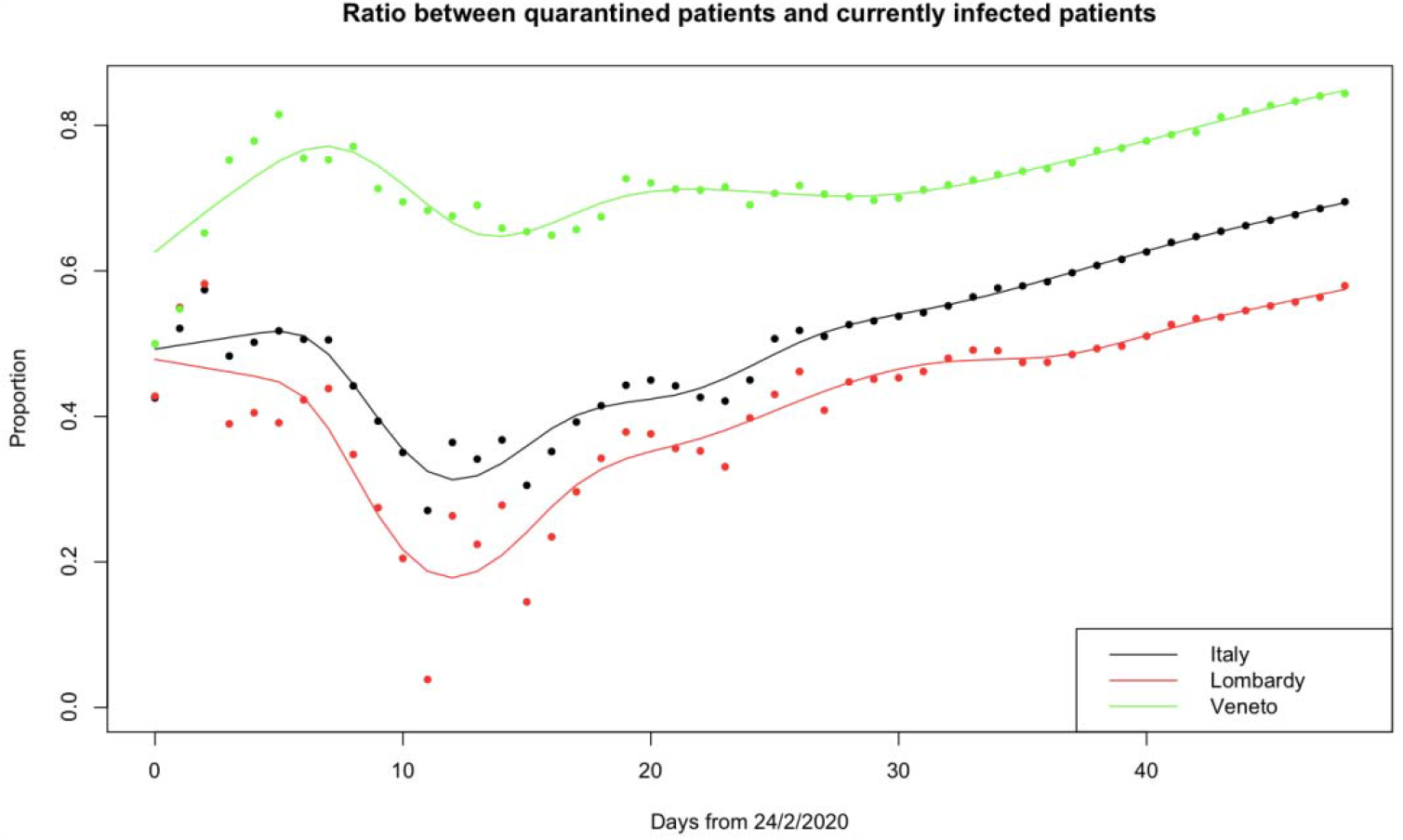
Ratio between quarantined patients and currently infected patients

The trend shows the strategy of care for this epidemic in Veneto whose government decided to bring to hospitals just the more severe patients, while treating the other ones at home.

So, we have two different regional strategies: the stable trend of Veneto is the result of the territorial care and the lower proportion of the Lombardy is the result of the hospital-centered care in this region (the initial fall followed by an increase is the result of the saturation of bed in hospital, particularly in ICUs).

Fig. 6 shows the ratio between symptomatic hospitalized patients and the total number of cases currently infected patients

**Fig. 6.**
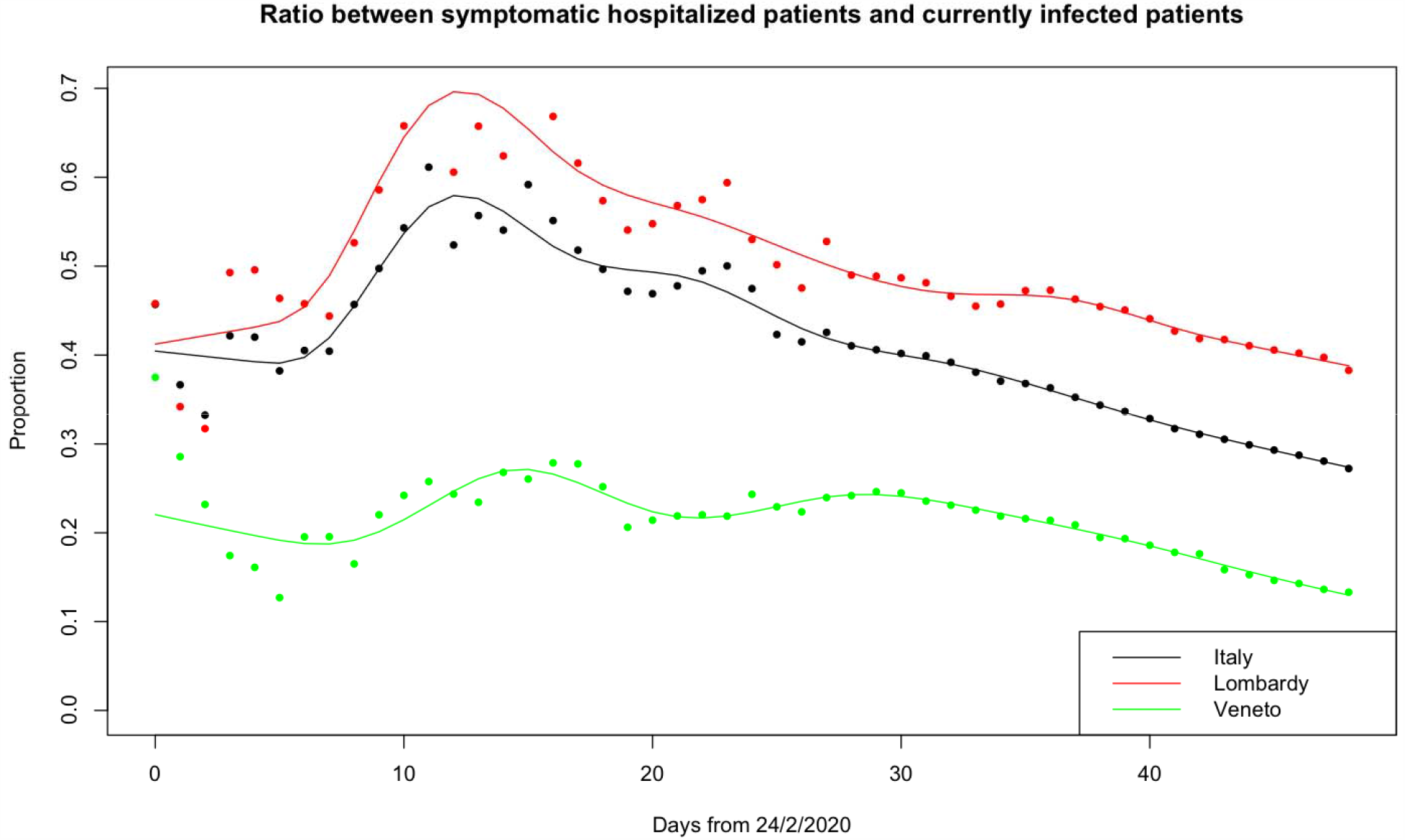
Ratio between symptomatic hospitalized patients and the total number of cases currently infected patients

This figure shows how in Lombardy the only strategy played for positive patients was the admission to hospital. The Veneto strategy seems to be more stable during all the period of observation.

However, the excess of positive cases related to early diagnosis can only partly explain the difference in apparent lethality due to the presence of silent cases without Covid-19 disease manifestation after massive screening practice in Veneto with respect to the symptomatic confirmation approach adopted in Lombardy. It is well known that this paradoxical effect is partly compensated by the increase of the lethality ratio in time even for the best regions. However, this phenomenon was observed also to the same extent for both Lombardy and Veneto. Better indicators should be therefore the ratio between the deaths and people hospitalized. In a region with lower degree of hospitalization we could expect that more serious cases are hospitalized therefore we could expect a higher ratio between the total number of deaths and the total number of hospitalized patients. This index should be at least comparable between the regions even in situations with different numbers of silent asymptomatic cases if similar admission criteria were adopted according to the similar expression of Covid-19 disease. If Veneto was expecting to admit more serious cases mostly relying for the rest on home management such an index was expected to be higher than Lombardy. However, the approximated index was 0.554 for Veneto and 0.737 for Lombardy (since the lack of analytical epidemiological data, the total number of hospitalized is only approximated by the sum of the presently hospitalized patients with the fraction of the total dismissed/healed proportional to the number of the hospitalized patients themselves). This result is supporting the fact that not only the apparent but also the true lethality could be also different between the two regions according to the different impact of Covid-19 on the local HS.

This evidence is also supported by the general mortality data and in particular to the relative mortality estimation looking at two reference cities of Lombardy: Milano and Brescia which were showing a huge increment in the expected mortality at 1.96 and 3.15 times with 1369 and 391 excess deaths, respectively, compared to Venezia and Verona at 1.16 and 1.33 times with 62 and 87 excess deaths, respectively attributable as directed and indirected causes to Covid-19 (8).

## Discussion

Looking at all these data it is possible to argue that the HS in Lombardy seems less prepared to cope with this epidemic while the Veneto HS managed quite well the patients and their disease.

These two regions represent the Italian situation where many different health organizations coexist: in Lombardy there was a progressive removal of public services in favor of private ones and there was the gradual dismantling of “territorial-centered” services and interventions (for example GPs, Local Social and Health Agencies) versus “hospital-centered” ones (for example the Emergency Departments); in Veneto we had a “territory-centered” system (9).

These features are reflected in the different health policies adopted by the two local governances to face CoViD-2019 epidemic.

In all the figures (except for a short trait in the ICUs pts. vs. currently infected pts. ratio curve), the national curve (which obviously is the result of a weighted average of all regional ones) lays between the two curves of Lombardy and Veneto, which are two “opposite poles” in the Italian landscape. Indeed, while we can consider the last one as a positive coping region. In particular, in 2009, in Lombardy, the Regional Management Board asked to limit the use of hospitals only to the more severe cases (10,11).

To face epidemic like this one the answer should be outside hospitals but this means to have general practitioners able to perform swabs and provided with PPEs, and in general well-trained healthcare personnel working outside hospitals (12).

This is not the Italian situation in fact the Global Health Security Index 2019 gave to Italy a very low score for emergency preparedness and response planning and risk communication: Italy was 31st on 195 Countries: Italy must start from this point (13, 14).

## Data Availability

The data are open available from national Covid-19 dashboards and repositories

https://github.com/pcm-dpc/COVID-19

## Author Contributions

SC, EB and BAR contributed conception and design of the study. BAR and EL organized the database. PB, EL and GM performed the statistical analysis and wrote the first draft of the manuscript. EP, LR, FA, FN, AM, GM and SS wrote sections of the manuscript SC, EB and MG reviewed and editing the manuscript. All authors contributed to manuscript revision, read, and approved the submitted version.

